# Comparison of Demographic and Diagnostic Markers in New Onset Pediatric Type 1 and Type 2 Diabetes

**DOI:** 10.1101/2021.08.16.21262068

**Authors:** Teresa Nieto, Beatriz Castillo, Jacobo Nieto, Maria J. Redondo

## Abstract

**Purpose:** Type 1 diabetes (T1D) is the most common type of diabetes in children, but the frequency of type 2 diabetes (T2D) is increasing rapidly. Classification of diabetes is based on a constellation of features that are typical of each type. We aimed to compare demographic, clinical and laboratory characteristics at diabetes diagnosis in pediatric T1D and T2D.

**Methods:** We studied children who attended a large academic hospital in Houston, Texas (USA) with a new diagnosis of T2D (n=753) or T1D (n=758). We compared age, sex, race/ethnicity, presence of obesity, glucose, hemoglobin A1c, islet autoantibody positivity, C-peptide, and presence of diabetic ketoacidosis (DKA) at diabetes diagnosis.

**Results:** At diagnosis of diabetes, children with T2D, compared with those with T1D, were older (13.6 vs 9.7% years old), more likely females (63.2% vs 47.8%), of racial/ethnic minority (91.1%% versus 42.3%) and obese (90.9% vs 19.4%), and were less likely to have DKA (7.8% vs 35.0%) and diabetes autoantibodies (5.5% vs 95.4%). Children with T2D also had significantly less marked elevation of glucose and hemoglobin A1c, and lower C-peptide levels (all comparisons, p<0.0001). In multiple logistic regression analysis, older age, racial/ethnic minority, obesity, higher C-peptide and negative islet autoantibodies were independently associated with T2D (all, p<0.05) while sex, glucose, hemoglobin A1c and DKA were not (model p<0.0001).

**Conclusions:** There are important demographic, clinical and laboratory differences between T1D and T2D in children. However, none of the characteristics was unique to either diabetes type, which poses challenges to diabetes classification at diagnosis.

## Introduction

Diabetes is the second most common chronic disease of childhood, after asthma. It occurs in 1 out of 300 children in the United States but its frequency is increasing, particularly among non-White racial groups (1). Most cases of pediatric diabetes are type 1 diabetes (T1D) (2), characterized by a lack of endogenous insulin that, in most cases, is due to autoimmunity against the beta-cells in the pancreatic islets. The appearance of islet autoantibodies in serum is a marker of the autoimmune process (3; 4). In contrast, in T2D, there is endogenous insulin secretion but not sufficient to meet the demands, which are frequently increased (insulin resistance) (3; 4). Insulin resistance is most often caused by obesity but is also promoted by puberty, pregnancy, aging and certain drugs. The “obesity epidemic” has caused a rise in T2D and it is now diagnosed in over 10% of children with diabetes (5).

Classification of diabetes type is currently based on the presence of a constellation of features that are typical of each diabetes type (3; 4). Of note, these characteristics were defined based on studies conducted in primarily non-Hispanic White populations, which may not be generalizable to other racial or ethnic groups. In addition, T2D is a relatively new cause of diabetes in children and the timely and correct identification of diabetes type is often challenging (6; 7). Therefore, we aimed to compare the characteristics at diabetes diagnosis of racially/ethnically diverse children with T1D or T2D.

## Materials and Methods

This is a secondary analysis of deidentified data from chart reviews of children newly diagnosed with T1D (n=758) between 2008 and 2010 (8) or T2D (n=753) between 2016 and 2019 (9) at a large academic hospital in Houston, Texas. Diabetes type was clinically assigned by their pediatric endocrinologist. These studies were approved by the Baylor College of Medicine Institutional Review Board (IRB).

We analyzed age, sex, race/ethnicity, presence of obesity, glucose, hemoglobin A1c, random C-peptide (a measure of endogenous insulin secretion (10)), presence of diabetic ketoacidosis (DKA) and positivity for islet autoantibodies (to insulin, GAD65 or IA-2/ICA512). All the variables analyzed were measured at diagnosis of diabetes. To describe each of the cohorts we used proportions for categorial variables and mean and standard deviation for continuous variables. To compare characteristics, we used Chi-square for categorial variables and t-tests for continuous variables. Multiple logistic regression analysis was used to examine the association between variables and type 2 diabetes with adjustment for potential confounders. To dichotomize continuous variables, we used the mean value of the distribution in the combined cohort, rounded to the closest whole number. All analyses were performed using STATA12 (StataCorp. 2011. *Stata Statistical Software: Release 12*. College Station, TX: StataCorp LP). Statistical significance was noted if 2-sided p-values were <0.05.

## Results

Children with T2D, compared with those with T1D, were significantly older (13.6 vs 9.7 years old) (p<0.0001) (Table 1). Diagnosis at 12 years old or greater occurred in 74.8% of the children with T2D compared with 31.3% of those with T1D (p<0.0001) (Figure 1). T2D was significantly associated with female gender (63.2% vs 47.8%, p<0.0001), minority race/ethnicity (91.1%% versus 42.3%, p<0.0001) and obesity (90.9% vs 19.4%, p<0.0001). Children with T2D had significantly lower glucose (247.7 mg/dl) than those with T1D (402.9 mg/dl, p<0.0001); glucose was under 300 mg/dl in 75.5% of the children with T2D and 32.8% of those with T1D (p<0.0001). Hemoglobin A1c was also lower in T2D (9.6%) than T1D (11.7%, p<0.0001); values under 11% were found in 67.6% of children with T2D and 35.5% of those with T1D (p<0.0001). C-peptide was higher in T2D (4.0 ng/ml) than T1D (0.65 ng/ml, p<0.0001); values of 2 ng/ml or above were found in 71.6% of the children with T2D and 4.6% of those with T1D (p<0.0001). DKA was significantly less frequent in T2D (7.8%) than in T1D (35.0%, p<0.0001). Most (95.4%) of children with T2D but only 5.5% of those with T1D were negative for islet autoantibodies (p<0.0001).

**Table 1.** Comparison of characteristics at diagnosis of T2D and T1D in children

| **Characteristic** | **T1D**<br>**N=758** | **T2D**<br>**N=753** | **p-value** |
| --- | --- | --- | --- |
| **Age (years)** | 9.7 (4.1)<br>Range: 0.66-18.1 | 13.6 (2.4)<br>Range: 6.0-18.9 | **<0.0001** |
| **Sex** |  |  | **<0.0001** |
| Female | 47.8% | 63.2% |  |
| Male | 52.2% | 36.8% |  |
| **Race/ethnicity** |  |  | **<0.0001** |
| Non-Hispanic White | 57.7% | 8.9% |  |
| Hispanic | 21.3% | 59.1% |  |
| Black | 16.3% | 28.9% |  |
| Other | 4.7% | 3.1% |  |
| **Obesity** | 19.4% | 90.9% | **<0.0001** |
| **Glucose (mg/dl)** | 402.9 (200.3)<br>Range: 63-1967 | 247.7 (136.6)<br>Range: 66-1213 | **<0.0001** |
| **Hemoglobin A1c (%)** | 11.7 (2.2)<br>Range: 5-15 | 9.6 (2.7)<br>Range: 5.1-15 | **<0.0001** |
| **C-peptide (ng/ml)** | 0.65 (0.9)<br>Range: 0.04-18 | 4.0 (3.6)<br>Range: 0.12-45.9 | **<0.0001** |
| **DKA** | 35.0%% | 7.8% | **<0.0001** |
| **Diabetes autoantibodies** |  |  | **<0.0001** |
| All three negative | 5.5% | 95.4% |  |
| >=1 positive | 94.5% | 4.6% |  |

Values are mean (SD) except where noted. Missing data: T1D: Age: n=1, sex: n=0, race/ethnicity: n=20, obesity: n=104, glucose: n=13, hemoglobin A1c: n=25, C-peptide: n=39, DKA: n=26, diabetes autoantibody positivity: n=13. T2D: Age: n=0, sex: n=0, race/ethnicity: n=15, obesity: n=15, glucose: n=103, hemoglobin A1c: n=90, C-peptide: n=260, DKA: n=11, diabetes autoantibody positivity: n=162. Abbreviations: T1D: Type 1 diabetes; T2D: Type 2 diabetes; DKA: Diabetic ketoacidosis

**Figure 1.**
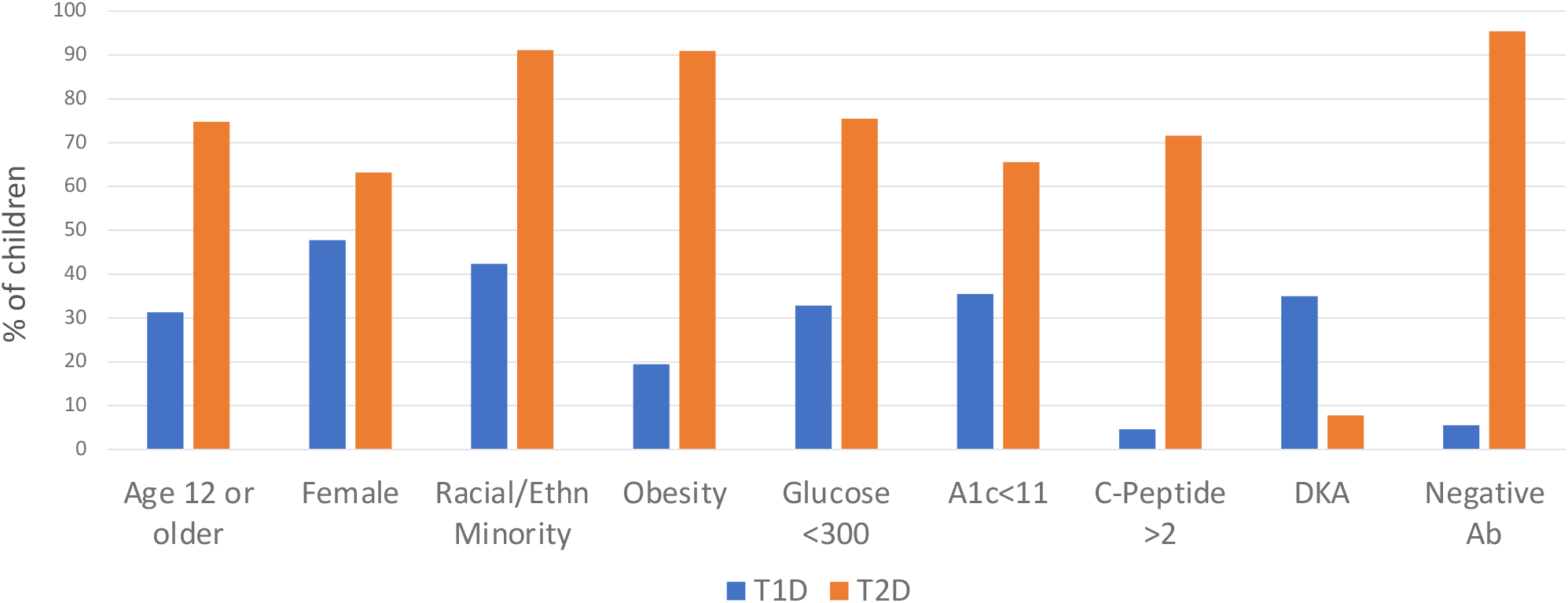
Comparison of characteristics in children with T1D and T2D at diagnosis (all, p<0.0001). Continuous variables were dichotomized using the mean value of the distribution in the combined cohort, rounded to the closest whole number. Abbreviations: T1D: Type 1 Diabetes; T2D: Type 2 Diabetes. A1c: Hemoglobin A1c. DKA: Diabetic ketoacidosis. Ab: Islet autoantibodies

In multivariable logistic analysis model (p<0.0001, n=1,020), T2D was significantly associated with older age (p=0.024), race/ethnicity (Hispanic ethnicity and African American race, versus non-Hispanic White, respectively, p=0.003 and 0.032), obesity (p<0.0001), higher C-peptide (p<0.0001) and negative islet autoantibodies (p<0.0001), but not with DKA (p=0.060), hemoglobin A1c (p=0.164), gender (p=0.253) or glucose (p=0.149) (Table 2).

**Table 2.** Multiple logistic regression analysis of T2D in children (n=1,023; p<0.0001)

|  | Odds Ratio | Std. Err | 95% Conf. Interval | P-value |
| --- | --- | --- | --- | --- |
| Age at diagnosis | 1.21 | .102 | 1.025-1.428 | **0.024** |
| Race/ethnicity |  |  |  |  |
| Non-Hispanic White | - | - | - | - |
| Hispanic | 5.66 | 3.351 | 1.773-18.059 | **0.003** |
| African American | 3.94 | 2.523 | 1.121-13.827 | **0.032** |
| Other | 2.48 | 2.45 | 0.358-17.140 | 0.358 |
| Obesity | 29.79 | 15.833 | 10.510-84.428 | **0.0001** |
| C-peptide | 1.96 | 0.374 | 1.347-2.847 | **0.0001** |
| Autoantibody positivity | 0.004 | 0.002 | 0.001-0.011 | **0.0001** |
| DKA | 0.32 | 0.194 | 0.099-1.049 | 0.060 |
| Hemoglobin A1c | 0.87 | 0.089 | 0.708-1.060 | 0.164 |
| Glucose | 0.998 | 0.001 | 0.995-1.000 | 0.149 |
| Gender | 1.679 | 0.761 | 0.691-4.083 | 0.253 |

## Discussion

We studied 758 children with T1D and 753 children with T2D at the time of diabetes diagnosis. Children with T2D were older and more likely to be of racial/ethnic minority and obese, less likely to have positive islet autoantibodies, and had higher C-peptide than children with T1D. These typical characteristics for T1D or T2D are currently used to classify diabetes type at onset (3) (4).

However, we also observed overlap in most of these characteristics. Although the mean age at diagnosis of T1D was significantly lower than in T2D, there was marked overlap in ranges of age at diagnosis (Table 1). T2D was diagnosed at age 12 or above in 74.8% of the children with T2D but 25.2% were younger. A recent study has reported children diagnosed with T2D under age 10, although it is still very uncommon (9).

Racial and ethnic distribution was different between T1D and T2D, and racial/ethnic minority was significantly associated with T2D, consistently with previously published reports (7). However, almost 9% of the children with T2D were non-Hispanic White and as much as 42.3% with T1D were of racial/ethnic minority. Obesity was also associated with T2D but was absent in 9.1% of the cases and was present in almost 20% of children with T1D. Therefore, race, ethnicity or obesity cannot be used to rule out a specific diabetes type.

Glucose and hemoglobin A1c were lower in T2D than in T1D but there was marked overlap in the range found at diagnosis in children with T1D or T2D. These characteristics were not significantly associated with diabetes type in the multivariable analysis. In sum, glucose and hemoglobin A1c alone cannot be used to differentiate diabetes type.

C-peptide was higher in the children with T2D than in those with T1D and the difference remained after adjustment for potential confounders. However, there was overlap between the ranges in the two groups (0.12-45.9 ng/ml in T2D and 0.04-18 ng/ml in T1D). C-peptide of 2 ng/ml or greater was found in most (71.6%) but not all of the children with T2D, and also in 4.6% of those with T1D. Therefore, C-peptide has limitations to as criterion for diabetes classification.

DKA is less common than in the past but still seen in 30-40% children with T1D at presentation (11). In our study, 35.0% of the children with T1D had DKA at diagnosis. Although much less common in T2D, we observed it in 7.8% of the children, which is consistently with recent data (12). Of note, with adjustment for the characteristics in the multivariable regression model (Table 2), DKA was not significantly associated with diabetes type. Overall, it is clear that DKA cannot be use to classify diabetes.

Islet autoantibodies were present in the vast majority (95.4%) of the children with T1D but also in 5.5% of the children with T2D. Potential reasons for the absence of autoantibodies in individuals with otherwise undistinguishable T1D could be the expression of autoantibodies that are not currently measured (13) or unrecognized forms of diabetes (14). Positivity for islet autoantibodies in a small percentage of children with clinically diagnosed T2D has been previously reported although its significance is still unclear (8; 15). These findings limit the use of islet autoantibody positivity to discriminate between diabetes types.

Our study had limitations. We were only able to study clinical characteristics at the time of diabetes diagnosis. Future studies will include genetic data and longitudinal follow-up. Important strengths of our study were the large sample size for both T1D and T2D, and the extensive clinical characterization.

In conclusion, we have described demographic, clinical, and laboratory features of children with T2D that are different from those of children with T1D, at diagnosis of their diabetes. However, the overlap in most of the typical features for each type may pose a challenge for clinicians to classify diabetes correctly and timely.

## Data Availability

Data are available upon reasonable request.

## Abbreviations

T1D: Type 1 diabetes
T2D: type 2 diabetes
DKA: Diabetic ketoacidosis

## Acknowledgements

The T1D cohort was collected with funds from the Texas Hospital Children Pilot Award (year 2010) (MJR). We acknowledge Dr. Marcela Astudillo, who collected the T2D cohort and provided feedback on the analyses, and Dr. Mustafa Tosur who also provided feedback on the analyses. The authors have no relevant conflict of interest to disclose.

## Author Contributions

T.N. contributed to statistical analyses and wrote the first draft of the manuscript. B.C. and J.N. contributed to data interpretation and revised/edited the manuscript. M.J.R. designed the study, supervised the statistical analyses and revised/edited the manuscript. MJR is the guarantor of this article and takes full responsibility for the work as a whole, including the study design, access to data, and the decision to submit and publish the manuscript.

## Data availability statement

Data are available upon reasonable request.

## References

1. Mayer-Davis EJ, Lawrence JM, Dabelea D, Divers J, Isom S, Dolan L, Imperatore G, Linder B, Marcovina S, Pettitt DJ, Pihoker C, Saydah S, Wagenknecht L, Study SfDiY. Incidence Trends of Type 1 and Type 2 Diabetes among Youths, 2002-2012. N Engl J Med 2017;376:1419–1429

2. Pettitt DJ, Talton J, Dabelea D, Divers J, Imperatore G, Lawrence JM, Liese AD, Linder B, Mayer-Davis EJ, Pihoker C, Saydah SH, Standiford DA, Hamman RF, Group SfDiYS. Prevalence of diabetes in U.S. youth in 2009: the SEARCH for diabetes in youth study. Diabetes Care 2014;37:402–408

3. American Diabetes Association. 2. Classification and Diagnosis of Diabetes:. Diabetes Care 2021;44:S15–S33

4. Mayer-Davis EJ, Kahkoska AR, Jefferies C, Dabelea D, Balde N, Gong CX, Aschner P, Craig ME. ISPAD Clinical Practice Consensus Guidelines 2018: Definition, epidemiology, and classification of diabetes in children and adolescents. Pediatr Diabetes 2018;19 Suppl 27:7–19

5. Liese AD, D’Agostino RB, Hamman RF, Kilgo PD, Lawrence JM, Liu LL, Loots B, Linder B, Marcovina S, Rodriguez B, Standiford D, Williams DE, Group SfDiYS. The burden of diabetes mellitus among US youth: prevalence estimates from the SEARCH for Diabetes in Youth Study. Pediatrics 2006;118:1510–1518

6. Jensen ET, Dabelea D. Type 2 Diabetes in Youth: New Lessons from the SEARCH Study. Curr Diab Rep 2018;18:36

7. Siller AF, Tosur M, Relan S, Astudillo M, McKay S, Dabelea D, Redondo MJ. Challenges in the diagnosis of diabetes type in pediatrics. Pediatr Diabetes 2020;

8. Redondo MJ, Rodriguez LM, Escalante M, Smith EO, Balasubramanyam A, Haymond MW. Types of pediatric diabetes mellitus defined by anti-islet autoimmunity and random C-peptide at diagnosis. Pediatr Diabetes 2013;14:333–340

9. Astudillo M, Tosur M, Castillo B, Rafaey A, Siller AF, Nieto J, Sisley S, McKay S, Nella AA, Balasubramanyam A, Bacha F, Redondo MJ. Type 2 Diabetes in Prepubertal Children. Pediatr Diabetes 2021 (in press)

10. Horwitz DL, Kuzuya H, Rubenstein AH. Circulating serum C-peptide. A brief review of diagnostic implications. N Engl J Med 1976;295:207–209

11. Rewers A, Klingensmith G, Davis C, Petitti DB, Pihoker C, Rodriguez B, Schwartz ID, Imperatore G, Williams D, Dolan LM, Dabelea D. Presence of diabetic ketoacidosis at diagnosis of diabetes mellitus in youth: the Search for Diabetes in Youth Study. Pediatrics 2008;121:e1258–1266

12. Praveen PA, Hockett CW, Ong TC, Amutha A, Isom SP, Jensen ET, Mohan V, Dabelea DA, D’Agostino RB, Hamman RF, Mayer-Davis EJ, Lawrence JM, Dolan LM, Kahn MG, Madhu SV, Tandon N. Diabetic ketoacidosis at diagnosis among youth with type 1 and type 2 diabetes: Results from SEARCH (United States) and YDR (India) registries. Pediatr Diabetes 2021;22:40–46

13. Acevedo-Calado MJ, Pietropaolo SL, Morran MP, Schnell S, Vonberg AD, Verge CF, Gianani R, Becker DJ, Huang S, Greenbaum CJ, Yu L, Davidson HW, Michels AW, Rich SS, Pietropaolo M, Group TDTS. Autoantibodies Directed Toward a Novel IA-2 Variant Protein Enhance Prediction of Type 1 Diabetes. Diabetes 2019;68:1819–1829

14. Urrutia I, Martínez R, Rica I, Martínez de LaPiscina I, García-Castaño A, Aguayo A, Calvo B, Castaño L, Group SPDC. Negative autoimmunity in a Spanish pediatric cohort suspected of type 1 diabetes, could it be monogenic diabetes? PLoS One 2019;14:e0220634

15. Klingensmith GJ, Pyle L, Arslanian S, Copeland KC, Cuttler L, Kaufman F, Laffel L, Marcovina S, Tollefsen SE, Weinstock RS, Linder B, Group TS. The presence of GAD and IA-2 antibodies in youth with a type 2 diabetes phenotype: results from the TODAY study. Diabetes Care 2010;33:1970–1975

